# Hyperpyrexia leading to death in a patient with severe COVID-19 disease

**DOI:** 10.1101/2020.05.18.20097220

**Authors:** Tanu Singhal, Sourabh Phadtare, Sunil Pai, Amit Raodeo

**Affiliations:** Department of Infectious Disease, Kokilaben Dhirubhai Ambani Hospital and Medical Research Institute, Mumbai, India; Department of Critical Care Medicine, Kokilaben Dhirubhai Ambani Hospital and Medical Research Institute, Mumbai, India

**Author notes:** **Corresponding author** Tanu Singhal, Consultant Infectious Disease, Kokilaben Dhirubhai Ambani Hospital and Medical Research Institute, Mumbai, India.

## Abstract

We describe here the clinical course of a 42 year old male with severe COVID 19 disease treated at a private hospital in Mumbai, India. This patient with very high inflammatory markers at admission was treated with supportive care, mechanical ventilation, anticoagulation, hydroxychloroquine, corticosteroids, tocilizumab, intravenous insulin, antibiotics, sedation and paralysis. There was sustained improvement in his respiratory status and decline in ventilator settings with decline and normalization of CRP, D dimer and PCT. However high fever persisted that did not respond to paracetamol and NSAIDS. On day 8 of admission his axillary temperature touched 107F followed by rapid clinical deterioration and death within the next 12 hours, Blood cultures were consistently sterile. While death was related to hyperpyrexia, the cause of this hyperpyrexia is uncertain.

## Case report

As the COVID-19 pandemic is evolving, new and atypical manifestations of the disease are being described regularly (1,2,3). We report a case of a 42 year old male with COVID-19 who was admitted to the COVID ICU of a tertiary care private hospital in Mumbai, India who had persistent fever and then hyperpyrexia leading to death. The Institutional Ethics and Research Committee gave a waiver for ethical approval for publication of this data.

This was a 42 year old male with normal body mass index (BMI) and who also had diabetes for past 8 years controlled on oral medications. He had symptoms of fever, cough and myalgia and after 6 days of symptoms tested positive for COVID-19 by RT PCR in the oropharyngeal swab. He was admitted to a local hospital on day 8 of illness and give symptomatic and supportive care. However he soon became breathless and was transferred to the study site on day 10 of illness. At admission he was drowsy, not obeying commands and tachypneic. His blood pressure was 146/62 mm of Hg and respiratory rate was 42/ minute and saturation on oxygen by mask was 63%. He was intubated and mechanically ventilated at arrival. His initial investigations revealed a Hb of 13.1, TLC of 10,720/ μl with an absolute lymphocyte count of 400 and ANC:ALC ratio of 25. The platelet count was 303000/ mm^3^. The serum CRP was 20 mg/dl with a procalcitonin of 1.5 ng/ml. The serum LDH 645 U/Land CPK was 308 U/L. The D dimer levels were more than 10000 FEU ng/ml. The serum creatinine was normal at 0.85 mg/dl. The liver function tests and coagulation profile (PT/ PTT) were normal. Blood cultures were sent. CXR showed bilateral infiltrates. Treatment was initiated with piperacillin tazobactam, azithromycin, hydroxychloroquine and methylprednisolone 30 mg twice daily. The patient was sedated with fentanyl and midazolam and paralysed with atracurium. On Day 0 of hospitalization the patient was also given tocilizumab 400 mg two doses 24 hours apart. Therapeutic 2 anticoagulation with enoxaparin 60 mg twice daily was also initiated. Intravenous insulin was given to control blood sugars. Over the next 2 days the patient needed high ventilator settings with PEEP of 12 cm of H_2_O, FiO_2_ of 70% with which the arterial PaO_2_ remained around 60 mm of Hg. After 48-72 hours there was decline in his ventilator requirements to a FiO_2_ of 40% and a PEEP of 6 with PaO_2_ of more than 100 mm of Hg. During the next 5 days there was a steady decline in inflammatory markers including CRP, D-Dimer, LDH and procalcitonin (Table 1) and rise in absolute lymphocyte counts and platelet counts. However the patient continued to run high fever ranging from 101-102°F. This fever did not respond to intravenous paracetamol. The blood and urine cultures were sterile and there was no other clinical focus of infection. Trial of ibuprofen, naproxen and even colchicine was given with no effect. External cooling measures were employed with which there was transient improvement. This fever was associated with tachycardia but there was no hypotension, oliguria or lactic acidosis. However on day 7, the fever touched 107°F. This was associated with high IL-6 levels of 220, high CPK of 12746 U/l and high ferritin levels of 7171 ng/ml but negative CRP and PCT. At this time the systolic BP dropped to 70 mm of Hg. Aggressive cooling with cold saline gastric lavage, fan, cooling blanket was initiated. Vasopressors were started, repeat blood cultures sent and empiric therapy with polymyxin B, meropenem and caspofungin was initiated. There was progressive downhill course with refractory hypotension, hyperthermia, oliguria, lactic acidosis and eventual cardiac arrest about 12 hours from the time the temperature touched 107°F.

**Table 1:** Laboratory parameters of the study patient.

| Day of hospitalization | 0 | 1 | 2 | 3 | 4 | 5 | 6 | 7 |
| --- | --- | --- | --- | --- | --- | --- | --- | --- |
| ALC /μl | 400 | 900 | 900 | 600 | 700 | 1200 | 1200 | 2500 |
| Platelets/μl | 303000 | 386000 | 427000 | 502000 | 500000 | 422000 | 417000 | 249000 |
| CRP (mg/dl) | 20 | 21.4 |  | 3.3 | 1.47 | 1 | 0.647 | 0.549 |
| PCT ( ng/ml) | 1.45 | 2.26 | 1.01 | 0.50 | 0.31 | 0.27 | 0.40 | 0.56 |
| LDH U/l | 645 | 434 |  |  |  |  | 471 | 720 |
| CPK U/l | 308 | 115 |  |  |  |  |  | 12746 |
| D Dimer FEU ng/ml | >10000 | 5914 | 3282 |  |  |  | 1155 | 809 |
| Serum creatinine mg/dl | 0.85 | 1.1 | 1.15 | 0.91 | 0.95 |  |  | 1.07 |
| LFT, RFT, coagulation profile | Normal |  |  |  |  |  |  |  |

ALC: absolute lymphocyte count, CRP: C reactive protein: PCT: Procalcitonin, LDH: Lactate dehydrogenase, CPK: creatinine phosphokinase, LFT: liver function tests

The non resolution of fever in the index case despite improving respiratory status, decline in inflammatory markers and no evidence of infection was puzzling. The most likely cause of this fever would be a cytokine storm especially since there was elevation of IL-6 and ferritin. But the other inflammatory markers including CRP and PCT were negative and D dimer had shown a sustained decline. Non response of this fever to NSAIDS and paracetamol suggested that fever could be due to temperature dysregulation (central fever). While there was no obvious neurologic deficit and the pupils were equal and reacting, it was not possible to assess the sensorium as patient was constantly sedated throughout is hospital stay. Neurologic involvement in COVID-19 has been described (4) The possibility of drug induced hyperthermia was also considered since the CPK was elevated on the day of hyperthermia. The urine myoglobin however was negative 825.9 μg/ml (cut off 1000 μg/ml) and the treatment chart review was negative for drugs causing hyperthermia (5). A PUBMED literature search using keywords“malignant hyperthermia” or “hyperpyrexia” and “COVID 19 did not yield any results.

While we are still not certain about the cause of hyperpyrexia, the temporal profile of events suggests that it was hyperpyrexia and resulting cellular dysfunction that caused death of this patient with COVID 19. This phenomenon should also be added to the already long list of atypical manifestations of COVID-19.

## Data Availability

The raw data of this case is available with the medical records department of the hospital

## References

1. Tay HS, Harwood R. Atypical presentation of COVID-19 in a frail older person [published online ahead of print, 2020 Apr 21]. Age Ageing. 2020;afaa068. doi:10.1093/ageing/afaa068

2. Kim J, Thomsen T, Sell N, Goldsmith AJ. Abdominal and testicular pain: An atypical presentation of COVID-19 [published online ahead of print, 2020 Mar 31]. Am J Emerg Med. 2020;S0735-6757(20)30194-7. doi:10.1016/j.ajem.2020.03.052

3. Ferrey AJ, Choi G, Hanna RM, et al. A Case of Novel Coronavirus Disease 19 in a Chronic Hemodialysis Patient Presenting with Gastroenteritis and Developing Severe Pulmonary Disease. Am J Nephrol. 2020;1–6. doi:10.1159/000507417.

4. Paniz-Mondolfi A, Bryce C, Grimes Z, et al. Central Nervous System Involvement by Severe Acute Respiratory Syndrome Coronavirus-2 (SARS-CoV-2). J Med Virol. 2020;10.1002/jmv.25915. doi:10.1002/jmv.25915

5. Walter E, Carraretto M. Drug-induced hyperthermia in critical care. J Intensive Care Soc. 2015; 16(4):306—311. doi:10.1177/1751143715583502

